# Mapping the Zone of Uncertainty in Pulmonary Function Test Interpretation

**DOI:** 10.1101/2025.06.03.25328441

**Authors:** Alexander T. Moffett, Meredith C. McCormack, Gary E. Weissman

## Abstract

**Background:** European Respiratory Society and American Thoracic Society (ERS/ATS) guidelines for pulmonary function test (PFT) interpretation posit the existence of a “zone of uncertainty.” However, as reference equations have been defined only for healthy lung function, the precise borders of the zone of uncertainty remain unspecified. To address this limitation, we sought to define distributions of both healthy and diseased lung function and to use these distributions to map the zone of uncertainty.

**Methods:** We used a latent class model to define distributions of healthy and diseased FEV_1_/FVC values. To represent the distribution of healthy FEV_1_/FVC values, we fit a Box-Cox Cole Green distribution to spirometry from healthy adult participants in the continuous National Health and Nutrition Examination Survey (NHANES). Using the distribution of healthy FEV_1_/FVC values, we then fit a latent class model to FEV_1_/FVC data from adult patients who underwent pulmonary function testing at the University of Pennsylvania Health System (UPHS) between 2000 and 2023. The distribution of diseased FEV_1_/FVC values and the prior probabilities that FEV_1_/FVC values had been sampled from healthy and diseased populations were selected to maximize the likelihood of the observed UPHS data. We considered the normal, Box-Cox Cole Green, and Box-Cox power exponential distributions for the distribution of diseased FEV_1_/FVC values and selected the distribution that minimized the Bayesian information criterion. We then mapped the zone of uncertainty by identifying the range of FEV_1_/FVC values in which the distributions of healthy and diseased lung function overlapped.

**Results:** Pre-bronchodilator spirometry data were collected from 14,075 NHANES participants— of whom 6,063 were without respiratory symptoms or a history of tobacco use—and 41,437 UPHS patients. Healthy lung function was represented by a Box-Cox Cole Green distribution with a median of 0.81, a coefficient of variation of 0.08, and skewness of 1.70. Diseased lung function was best represented by a Box-Cox power exponential distribution with a median of 0.56, a coefficient of variation of 0.30, skewness of 0.91, and kurtosis of 2.36. In the latent class model, the prior probability that an FEV_1_/FVC value was healthy was 76.2%, while the prior probability that it was diseased was 23.8%. The overlap of the distributions of healthy and diseased FEV_1_/FVC values defined a zone of uncertainty in the interval [0.64, 0.95]. The FEV_1_/FVC values of 33,747 (81.4%) UPHS patients fell within the zone of uncertainty, including 30,288 (99.2%) patients with a normal FEV_1_/FVC and 3,459 (31.7%) patients with an abnormal FEV_1_/FVC.

**Conclusion:** This exploratory study presents evidence that in a clinical cohort, most FEV_1_/FVC values fall within the zone of uncertainty. Further research is needed to develop optimal ways to represent and incorporate this uncertainty into PFT interpretation.

## Introduction

European Respiratory Society and American Thoracic Society (ERS/ATS) guidelines for pulmonary function test (PFT) interpretation posit the existence of a “zone of uncertainty” defined by the overlap of distributions of healthy and diseased lung function (**Figure 1a**).^1^ As PFT parameter values that fall within the zone of uncertainty may represent either health or disease, ERS/ATS guidelines recommend that these values be interpreted with caution.^1^,^2^ However, as reference equations have been defined only for healthy lung function, the precise borders of the zone of uncertainty remain unspecified. To address this limitation, we sought to define distributions of both healthy and diseased lung function and to use these distributions to map the zone of uncertainty.

**Figure 1.**
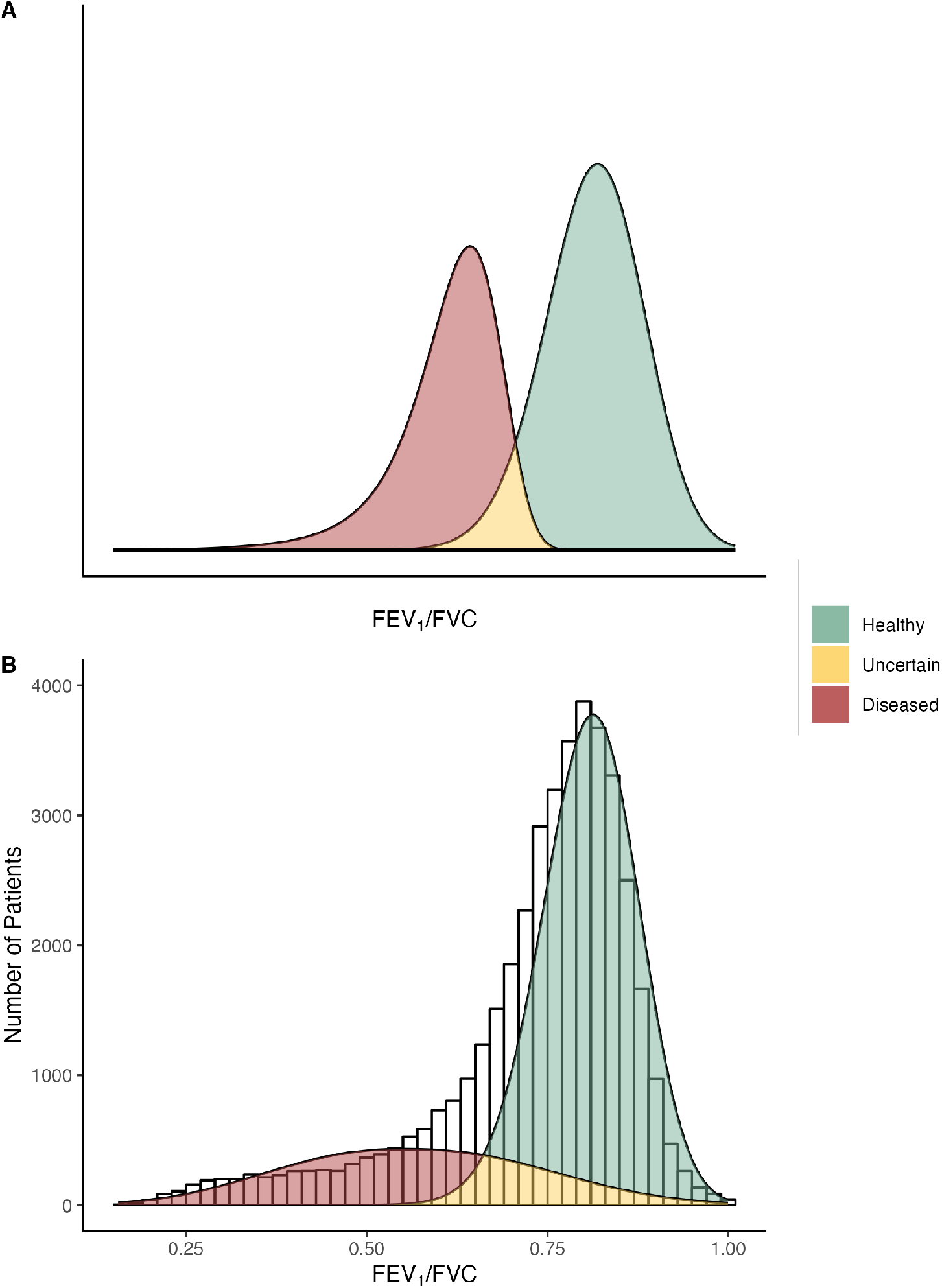
A latent class model of the FEV_1_/FVC in health and disease. (A) The theoretical distribution of the FEV_1_/FVC in health and disease, adapted from the “ERS/ATS Technical Standard on Interpretive Strategies for Routine Lung Function Tests,” Figure 5.^1^ The green and red distributions represent the FEV_1_/FVC in health and disease, while the yellow area, defined by the overlap of these distributions, represents the zone of uncertainty. (B) The empirical distribution of the FEV_1_/FVC in health and disease, from pulmonary function tests performed at three pulmonary diagnostic labs in the University of Pennsylvania Health System between 2000 and 2023 (*N* = 41,437). Observed FEV_1_/FVC values are binned by 0.02. A latent class model, defining distributions of healthy and diseased lung function, was fit to these data. The green and red probability distributions once again represent the FEV_1_/FVC in health and disease, while the yellow area, defined by the overlap of these distributions, represents the zone of uncertainty.

## Methods

We used a latent class model to define distributions of healthy and diseased lung function. In this exploratory study we considered only a single PFT parameter, the ratio of the pre-bronchodilator forced expiratory volume in 1 second to forced vital capacity (FEV_1_/FVC). To represent the distribution of healthy FEV_1_/FVC values, we fit a Box-Cox Cole Green distribution to spirometry from healthy adult participants in the continuous National Health and Nutrition Examination Survey (NHANES).^3^,^4^ We defined healthy participants as adults without respiratory symptoms or a history of tobacco use.^5^,^6^ Using the distribution of healthy FEV_1_/FVC values, we then fit a latent class model to FEV_1_/FVC data from adult patients who underwent pulmonary function testing at the University of Pennsylvania Health System (UPHS) between 2000 and 2023. In this model FEV_1_/FVC values were assumed to have been sampled from either a healthy or a diseased population, with the population from which a patient was sampled representing a latent class. In fitting this model, the distribution of diseased FEV_1_/FVC values and the prior probabilities that FEV_1_/FVC values had been sampled from healthy and diseased populations were selected to maximize the likelihood of the observed UPHS data.^7^ We considered the normal, Box-Cox Cole Green, and Box-Cox power exponential distributions for the distribution of diseased FEV_1_/FVC values and selected the distribution that minimized the Bayesian information criterion.^8^ We then mapped the zone of uncertainty by identifying the range of FEV_1_/FVC values in which the distributions of healthy and diseased lung function overlapped.

All code is open source and freely available (https://github.com/weissman-lab/zone). The University of Pennsylvania Institutional Review Board approved this study.

## Results

Pre-bronchodilator spirometry data were collected from 14,075 NHANES participants— of whom 6,063 were without respiratory symptoms or a history of tobacco use—and 41,437 UPHS patients. The median age of the UPHS patients was 57 (interquartile range [IQR] 21), with 23,753 (57.3%) women, and 27,058 (65.3%) non-Hispanic White patients. The median FEV_1_/FVC among the UPHS patients was 0.77 (IQR 0.14).

Healthy lung function was represented by a Box-Cox Cole Green distribution with a median of 0.81, a coefficient of variation of 0.08, and skewness of 1.70. The lower limit of normal (LLN), defined by a z-score of –1.645, was 0.069 for this distribution and 10,910 (26.3%) UPHS patients had an FEV_1_/FVC less than the LLN. Diseased lung function was best represented by a Box-Cox power exponential distribution with a median of 0.56, a coefficient of variation of 0.30, skewness of 0.91, and kurtosis of 2.36. In the latent class model, the prior probability that an FEV_1_/FVC value was healthy was 76.2%, while the prior probability that it was diseased was 23.8%.

The overlap of the distributions of healthy and diseased FEV_1_/FVC values defined a zone of uncertainty in the interval [0.64, 0.95] (**Figure 1b**). The FEV_1_/FVC values of 33,747 (81.4%) UPHS patients fell within the zone of uncertainty, including 30,288 (99.2%) patients with a normal FEV_1_/FVC and 3,459 (31.7%) patients with an abnormal FEV_1_/FVC.

## Discussion

Using a latent class model to define distributions of healthy and diseased lung function, we found that the overlap of these distributions—what ERS/ATS guidelines refer to as the zone of uncertainty—is much greater than has been assumed, encompassing almost all of the FEV1/FVC values in our clinical cohort.^9^ The FEV_1_/FVC values in our clinical cohort had a unimodal distribution, rather than the bimodal distribution assumed in ERS/ATS guidelines, and applying a latent class model to this unimodal distribution led to the substantial degree of overlap in our latent classes.

Our exploratory results suggest that uncertainty in the interpretation of FEV_1_/FVC values is pervasive. Such broad uncertainty may demonstrate a need to move beyond the binary classification of PFT interpretations as either certain or uncertain and instead develop methods to quantify the degree of uncertainty associated with such interpretation. One way to do so is to adopt a Bayesian approach to PFT interpretation, in which, rather than report whether an FEV_1_/FVC value falls above or below a lower limit of normal, we instead report the posterior probability that the value was sampled from a healthy population. The latent class model developed in this paper allows for the estimation of the parameters necessary to use Bayes’ theorem to calculate this posterior probability. While the appeal of a Bayesian approach to PFT interpretation is often noted, it has been seen by some to represent more of a theoretical ideal than a practical reality.^10–12^ Our results suggest the importance of adopting a Bayesian approach and provide an initial roadmap for how to do so.

Future efforts can address some of the limitations of our study. First, this exploratory study was limited to the FEV_1_/FVC. The FEV_1_/FVC was chosen as this is the parameter used by ERS/ATS guidelines to introduce the concept of the zone of uncertainty, and because it can be meaningfully interpreted independently of patient age, height, and sex, allowing for the construction of simple univariate models that can be easily visualized. However, as the FEV_1_/FVC is often elevated in patients with restrictive ventilatory impairments, this parameter may be less clearly delineated in health and disease than, for instance, the FEV_1_. Further work should consider other PFT parameters, while stratifying by sex and adding age and standing height as covariates. Second, as the distributions of healthy and diseased FEV_1_/FVC values assigned non-zero probabilities to all values in the interval [0,1], an additional threshold was needed to convert some of these probabilities to 0, so that the zone of uncertainty would not *a priori* encompass the entire interval. In this study we used the 0.01 and 0.99 quantiles to delimit these distributions. Further work should consider how other approaches might yield different depictions of the zone of uncertainty. Third, we considered only a subset of the many possible statistical distributions and clustering algorithms that could be used for latent class analysis. Further work assessing the sensitivity of these results to different modeling strategies is needed. Fourth, this is a single-site study and it is expected that distributions of diseased lung function will differ across different sites. To address this limitation, all code used in this study is freely available and open source, allowing for the development of analogous models for other clinical sites.

In conclusion, this exploratory study presents evidence that in a clinical cohort, most FEV_1_/FVC values fall within the zone of uncertainty. Further research is needed to develop optimal ways to represent and incorporate this uncertainty into PFT interpretation.

## Data Availability

All data produced in the present study are available upon reasonable request to the authors.

https://github.com/weissman-lab/zone

## Author Contributions

ATM participated in study design and analysis, drafted and revised the manuscript. MCM GEW participated in study design and revised the manuscript. All authors have read and approved the manuscript.

